# Evaluating transmission heterogeneity and super-spreading event of COVID-19 in a metropolis of China

**DOI:** 10.1101/2020.05.06.20073742

**Authors:** Yunjun Zhang, Yuying Li, Lu Wang, Mingyuan Li, Xiaohua Zhou

## Abstract

**Background:** COVID-19 caused rapid mass infection worldwide. Understanding its transmission characteristics including heterogeneity is of vital importance for prediction and intervention of future epidemics. In addition, transmission heterogeneity usually envokes super spreading events (SSEs) where certain individuals infect large numbers of secondary cases. Till now, studies of transmission heterogeneity of COVID-19 and its underlying reason are far from reaching an agreement.

**Methods:** We collected information of all infected cases between January 21 and February 26, 2020 from official public sources in Tianjin, a metropolis of China. . Utilizing a heterogeneous transmission model based on branching process along with a negative binomial offspring distribution, we estimated the reproductive number *R* and the dispersion parameter *k* which characterized the transmission potential and heterogeneity, respectively. Furthermore, we studied the SSE in Tianjin outbreak and evaluated the effect of control measures undertaken by local government based on the heterogeneous model.

**Results:** A total of 135 confirmed cases (including 34 imported cases and 101 local infections) in Tianjin by February 26th 2020 entered the study. We grouped them into 43 transmission chains with the largest chain of 45 cases and the longest chain of 4 generations. The estimated reproduction number *R* was at 0.67 (95%CI: 0.54~0.84), and the dispersion parameter *k* was at 0.25 (95% CI: 0.13~0.88). A super spreader causing six infections in Tianjin, was identified. In addition, our simulation results showed that the outbreak in Tianjin would have caused 165 infections and sustained for 7.56 generations on average if no control measures had been taken by local government since January 28th.

**Conclusions:** Our analysis suggested that the transmission of COVID-19 was subcritical but with significant heterogeneity and may incur SSE. More efforts are needed to verify the transmission heterogeneity of COVID-19 in other populations and its contributing factors, which is important for developing targeted measures to curb the pandemic.

## Background

In December 2019, many cases of viral pneumonia-like disease similar to severe acute respiratory syndrome (SARS) were detected in Wuhan, China. Later they were confirmed to be caused by a novel coronavirus, provisionally called 2019 novel coronavirus (2019-nCoV). On January 30th 2020, World Health Organization (WHO) declared the 2019-nCoV outbreak as a global health emergency of international concern [1]. On February 11th 2020, WHO named the diseases CoronaVirus Disease 2019 (COVID-19) and announced ”severe acute respiratory syndrome coronavirus 2 (SARS-CoV-2)” as the name of the new virus. On March 11th 2020, WHO declared COVID-19 a pandemic. As of April 18th 2020, more than 2,160,207 cases were confirmed globally and the National Health Commission (NHC) of China reported a total of 82,735 cases of COVID-19 in mainland China, including 77,062 recoveries and 4,632 deaths.

The dynamics of an infectious disease outbreak depends on both the potential and the heterogeneity of disease transmission. The transmission potential of an infectious pathogen is usually represented by its reproduction number (denoted as *R*) which is the average number of secondary cases caused by a typical infectious individual. Estimates of *R* > 1 (i.e., supercritical outbreak), such as for the early outbreak of the severe acute respiratory syndrome coronavirus (SARS-CoV) in China 2003 [2], indicate the great risk for an infection pathogen to generate a major outbreak; estimates of *R* < 1 (i.e., subcritical outbreak), such as for the Middle East respiratory syndrome coronavirus (MERS-CoV) in South Korea 2018 [3], imply that the outbreak is slowing down with declining trend of incidence. Though different in transmission potential, both SARS and MERS shared the same feature of high level of heterogeneity which involved the uneven transmission patterns with a number of super-spreading events (SSEs), where some individuals spread to a disproportionate number of individuals, as compared to most individuals who infected a few or none [2, 4]. SSEs of SARS and MERS were responsible for triggering the initial outbreaks in several large cities such as Beijing, Hongkong and Singapore, and hence sustaining the spread of disease worldwide. Besides, SSEs were also documented in many other infectious disease [5]. Detection of transmission heterogeneity may direct prevention efforts and reduce future infections [6, 7, 8].

The transmission potential of COVID-19 has been studied based on mathematical models, yielding consistent evidence for the high level of reproduction number *R* (1.95~2.2) within a completely susceptible population [9, 10, 11]. However, evidence for the transmission heterogeneity of COVID-19 and the related SSEs was limited and conflicting. So far, two studies found no heterogeneity in Singapore [12] and in a southern city of China [13], but one study confirmed heterogeneity based on data from 46 countries [14]. In addition, several possible SSEs have been reported in China[15, 16] which involved large numbers of infections. Hence, it would be essential to further explore the transmission heterogeneity and SSE in COVID-19 pandemic and its relevant factors

In this study, we investigated the transmission characteristics of COVID-19 using the epidemiological data of all the confirmed cases in a metropolis in Northern China. We analysed the transmission chains and then estimated the reproduction number and the transmission heterogeneity using a branching model along with a negative binomial offspring distribution [17]. In addition, we identified the SSE according to the 99-percentile criterion in [2] and assessed the effect of control measures employed by local government.

## Method

### Data Collection

Tianjin is a municipality and a coastal metropolis in Northern China, with a population of 15.66 million and an area of 11,966 square kilometers. Since the first case of COVID-19 in Tianjin was confirmed on January 21st, a total of 135 cases were confirmed by the time of data collection (February 26th, 2020) of the current study. From 6:00 on February 22 to 18:00 on February 26, the city had no new confirmed cases for 108 consecutive hours. The municipal government publicized the epidemiological information of the cases every day. Diagnosis of COVID-19 were based on the protocol issued by China CDC [18].

From the official websites of the Municipal Health Commissions [19], we retrieved data of the 135 confirmed COVID-19 cases in Tianjin, including demographic characteristics, epidemiological characteristics, i.e., travel history and contact history with confirmed/suspicious cases. According to these information, the infection relationship within the scope of Tianjin can be specified. Each case was given a unique number according to its sequential order as reported.

We adopted the definition in [17] to define a transmission chain as a group of cases connected by an unbroken series of local transmission events. We grouped all the confirmed cases in Tianjin into transmission chains and for each transmission chain, we identified the primary case, calculated the chain size (i.e., the total number cases including the primary case) and reconstructed the transmission history (i.e., who infected whom). According to the extent of resolution of the reconstructed information, three types of transmission chains were further identified. The first type was the *simple transmission chain* in which the transmission history could be clearly recovered. The second type was the *ordinary transmission chain* for which the transmission history was not clear but the single primary case could be clearly identified and the chain size was also clear. The third type was the *complex transmission chain* for which the chain size was clear but the primary case could not be clearly identified. More than one individual in the chain exhibited similar behavior/clinical characteristics, so we have to regard them as the primary cases.

As COVID-19 was first reported in Wuhan, a city in the middle of China, a primary case was defined if the individual had a history of travel to or residence in Wuhan within one month, had direct contact with an individual who had confirmed infection outside Tianjin, having fever outside Tianjin, or had the earliest onset of symptoms in the transmission chain.

### Analytical approach

#### Inference of Transmission Characteristics

To quantify the transmission potential and heterogeneity for the COVID-19 outbreak in Tianjin, we adopted a stochastic model based on branching process to characterize both the distribution of secondary cases (i.e., a negative binomial distribution) and the resulting distribution of transmission chain sizes under the same parameterization of reproduction number *R* and dispersion parameter *k* (lower value indicating higher heterogeneity) for subcritical epidemic [17].

We first fitted the stochastic model to the retrieved information of the simple/ordinary transmission chains. Then, in handling a complex transmission chain with two primary cases, we regarded it could be separated into two ordinary transmission chains, each of which was led by a primary case. The difficulty lay in that the exact size of each ordinary chain was unclear. We dealt with this ambiguity in two ways: one was the combinatorial method in [17] by allowing for all the possible combinations and treated the sum as an overall probability; the other was to adopt the expectation-maximization (EM) algorithm by treating the sizes of two separated chains as latent variables. The EM algorithm also estimated the latent chain size, which was informative for dividing the complex transmission chain into constituent chains (Section B, Appendix 1).

To identify the possible SSEs in Tianjin outbreak, we adopted the definition in [2] to define a super-spreader as any infected individual causing more infections than would occur in 99% of infectious histories in a homogeneous population. The transmission in a homogeneous situation was modelled with a *Poisson* distribution which is the special case of negative binomial distribution without heterogeneity.

#### Assessment of Control Measures

Based on the stochastic model accounting for transmission heterogeneity, we assessed the control measures imposed by the Tianjin government. We compared the transmission characteristics (i.e., *R* and *k*) for the periods of before and after the control measures taking effect. Then based on the estimate of transmission characteristics before control, we simulated the expected distribution of outbreaks that might occur in Tianjin if no control measures were taken. Each simulated outbreak was initiated with 43 infectious individuals (as the number of transmission chains in Tianjin), and was propagated with the branching process model on the basis of inferred characteristics.

## Results

### Characteristics of COVID-19 Cases

Among the 135 patients included in the study, 72 (53.3%) were males and 63 (46.7%) females, with an average (standard deviation) age of 47.8 (18.3) years and 50.1 (15.2) years, respectively. As shown by Fig.1 A, of the total 135 cases, 34 (25.2%) cases were imported cases. Among the 101 cases of local transmission, the majority (55, 40.7%) were infected in household, and 35 (25.9%) cases were infected in public places including a shopping mall and working places, and 11 (8.1%) were unclear in the source of infection. When we examined the chronological development of the infection in Tianjin, we found that the imported cases and the household infections dominated the confirmed cases of the early and the later stage of outbreak respectively. The transition happened around February 3rd.

**Figure 1:**
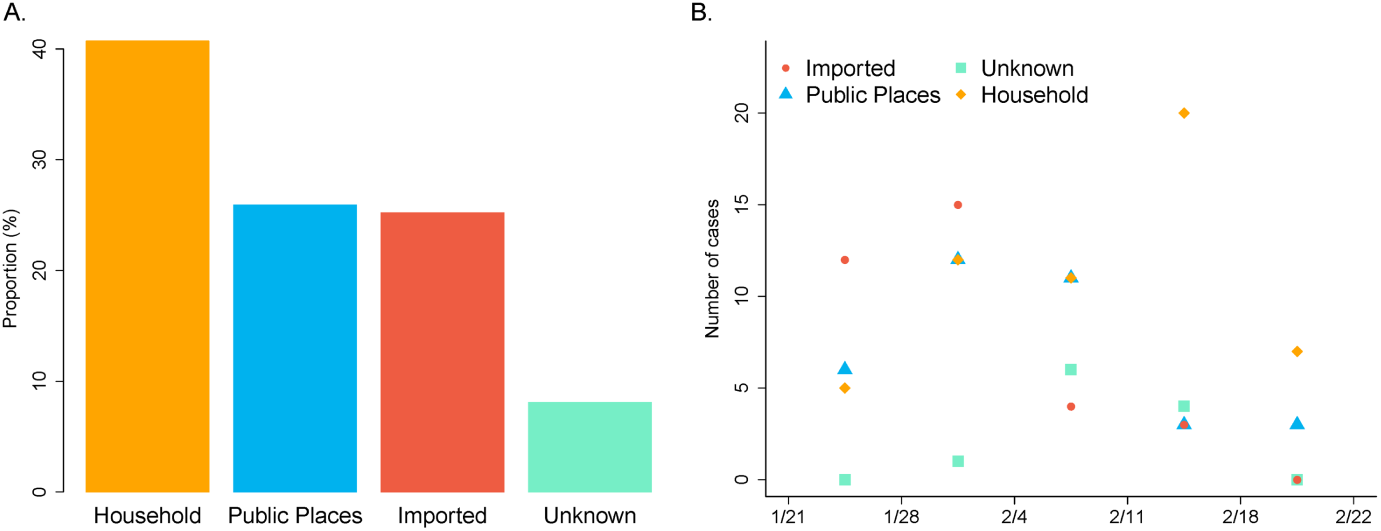
Case information in Tianjin (totally 135 cases, reported from January 21st to February 26th) A: Proportion of cases infected in different ways/places. B: Chronological development of the infection by transmission ways/places

### Reconstructed Transmission Chains

The 135 cases were grouped into 43 transmission chains including 36 simple chains (47 cases, average size 1.3), 5 ordinary chains (78 cases, average size 15.6), and 2 complex chains (10 cases, average size 5). (Table 1, Figure 2). Detail information of three type of transmission chains shown in Section A of additional file 1.

**Table 1:** Three types of COVID-19 transmission chains in Tianjin.

| Chain Type | Amount of Chains | Total Number of Cases | Average Chain Size | Range of Chain Size |
| --- | --- | --- | --- | --- |
| Simple transmission chain | 36 | 47 | 1.3 | 1 - 4 |
| Ordinary transmission chain | 5 | 78 | 15.6 | 3 - 45 |
| Complex transmission chain | 2 | 10 | 5 | 5 - 5 |

**Figure 2:**
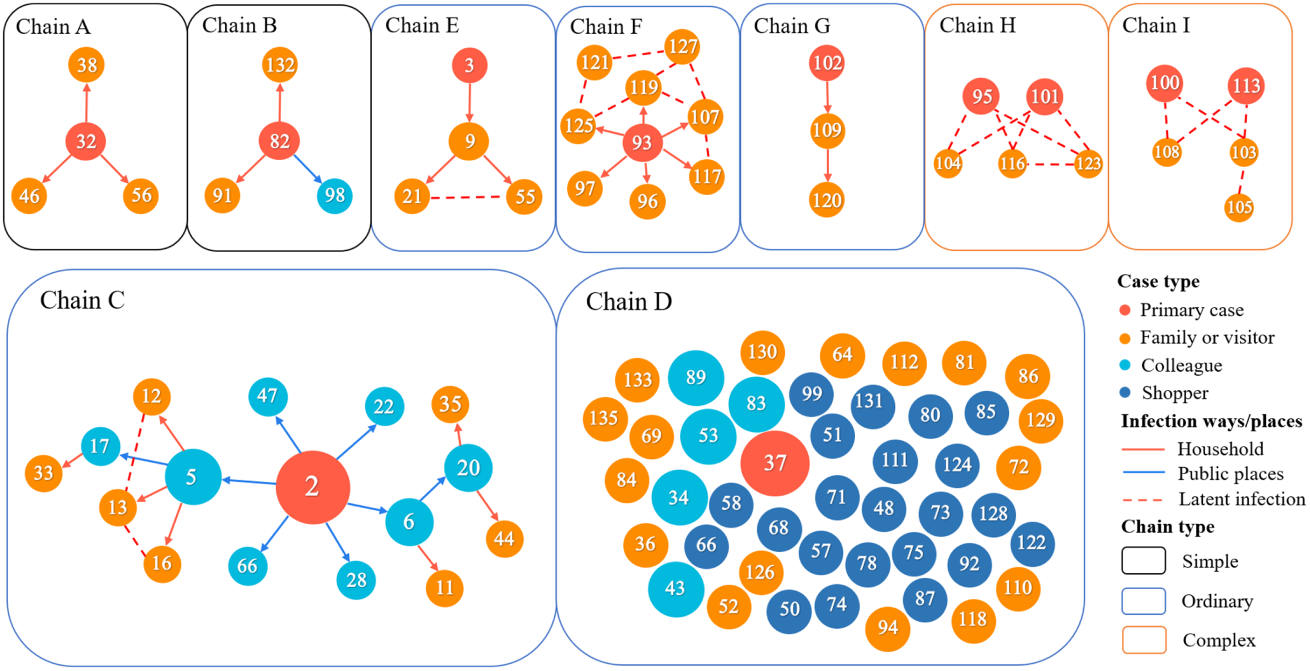
Reconstructed transmission chains (excepted for isolated cases) of COVID-19 outbreak in Tianjin by February 26th, 2020. The red circles represent the primary cases in each chain, the orange circles are the family members or visitors, and the blue circles are the colleagues. The red arrows and the blue arrows represent the transmissions within household and in public places respectively. The dash lines represent latent epidemiological links. The black/blue/orange frames represent the simple/ordinary/complex transmission chains respectively.

### Estimation of *R* and *k*

Both the combinational method and the EM algorithm gave lower value of *k* with the corresponding 95% confidence intervals (CIs) being lower than 1 (Table 2), suggesting significant evidence for the transmission heterogeneity in the outbreak at Tianjin. In addition, both combinatorial method and EM algorithm gave the same estimate of reproductive number *R* = 0.67 being lower than the critical value of 1, indicating the local transmission would finally die out. Note that these estimate of transmission potential was based on the data by February 26th, implying the average trend over the same period.

**Table 2:** Estimation and CI of the reproduction number *R* and dispersion parameter *k* based on the conbinational method and the EM algorithm

|  | Combinatorial method (95% CI) | EM (95% CI) |
| --- | --- | --- |
| $R$ | 0.67 (0.44, 1.03) | 0.67 (0.54, 0.84) |
| $k$ | 0.26 (0.10, 0.88) | 0.25 (0.13, 0.88) |

When comparing the CIs from different methods, we observed that the EM algorithm generated slightly narrower CI for both parameters than the combinatorial method. In addition, the EM algorithm had advantage on the ability of providing more information on the split of complex transmission chain since it also produced a probabilistic insight of unknown partitions of the chain. Recall that the COVID-19 data in Tianjin contains two complex transmission chains of size 5 for each. For these two complex chains, the EM algorithm tended to split it into one size-1 and one size-4 transmission chain (posterior probability = 95.5%), instead of two chains with size 2 and size 3 (posterior probability = 4.5%).

Furthermore, the estimates along with the 95% CI and confidence regions of combinatorial method were plotted in Fig. 3 A. The confidence region incorporated the uncertainty in both parameters simultaneously, thus it undoubtedly gave much wider range than the CI, especially for *k*. We also found (Fig. 3 B) that the observed chain size distribution was close to the expected distribution based on the estimated characteristics (i.e., 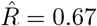 and 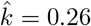). Particularly, the probability of a chain with size over 5 was less than 10%, and the probability of a single infected case resulting in a chain of size 45 (the largest size of transmission chain in Tianjin) was about 0.8%. It indicated that the outbreak of a comparatively large chain was not likely to happen in Tianjin.

**Figure 3:**
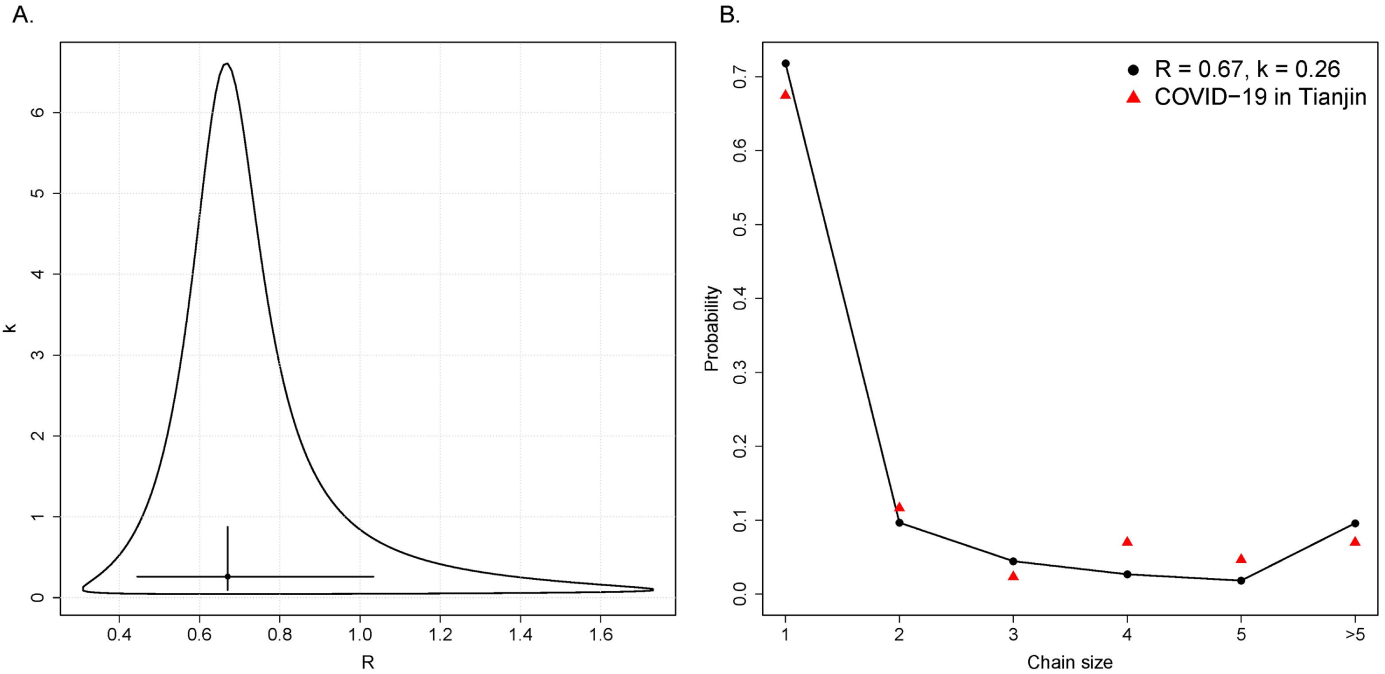
Analysis of COVID-19 outbreak in Tianjin using the combinatorial method. A: The circle, cross hair, and curve represent the estimates, 95% confidence intervals and confidence region of parameters *R* and *k*, respectively. B: Circles denote the probability of a transmission chain with size from 1 to > 5 based on the estimates of *R* and *k*; Triangle denotes the frequency of a transmission chain with corresponding size in Tianjin COVID-19 data.

### Super Spreading Event in Tianjin

We found notable transmission heterogeneity (*k* = 0.25) in the local spreading of COVID-19 in Tianjin, suggesting that there was likely to be SSEs [5]. As SSEs happened more likely at the early stage of the outbreak to trigger out the local spreading in the epidemic of SARS and MERS, we therefore had a reason to suspect the existence of SSEs at the early stage of outbreak in Tianjin.

Adopting the criterion in [2] to define SSE, we calculated the cut-off as the 99th percentile of the *Poisson* distribution with mean value of 2.2 (i.e., the reproduction number at the early), which was 6, suggesting that an infected individuals who infected 6 or more secondary cases could be regarded as a super spreader of the COVID-19 outbreak in Tianjin. According to this criterion, the second case confirmed in Tianjin was a super spreader. This primary case infected six colleagues within close contact before being confirmed on January 21st, 2020 (Fig.2). These six colleagues, as secondary cases in this transmission chain, successively infected other colleagues or their relatives.

### Effect of Government Control Measures

Tianjin municipal government deployed a series of policies to control the spread of COVID-19 from January 28th, 2020, including strong traffic restriction and quarantining individuals who had suspect contacts with confirmed cases[20]. By allowing for the median incubation period of 5.1 days [21], we assumed that the control measures had been taking effect since February 1st, 2020.

When we compared the transmission characteristics before and after the intervention taking effect (Table 3), we found that the reproduction number *R* decreased from 0.74 to 0.53, while *k* increased from 0.14 to 0.77, suggesting the decrease in both transmission potential and heterogeneity after taking control measures. However, the overlapping CIs were likely to be caused by the small sample size.

**Table 3:** Estimation of the reproductive number *R* and the dispersion parameter *k* for different periods

|  | Before February 1st (95% CI) | After February 1st (95% CI) |
| --- | --- | --- |
| $R$ | 0.74 (0.39, 1.61) | 0.53 (0.29, 0.96) |
| $k$ | 0.14 (0.04, 0.63) | 0.77 (0.14, 31.47) |

Compared with the observation of 135 confirmed cases in total and the longest transmission chain with 4 generations (Fig. 2), the simulation study showed that the local outbreak in Tianjin would have sustained for 7.56 generations and would have led to 165 infections on average if there were no control policies (Fig. 4).

**Figure 4:**
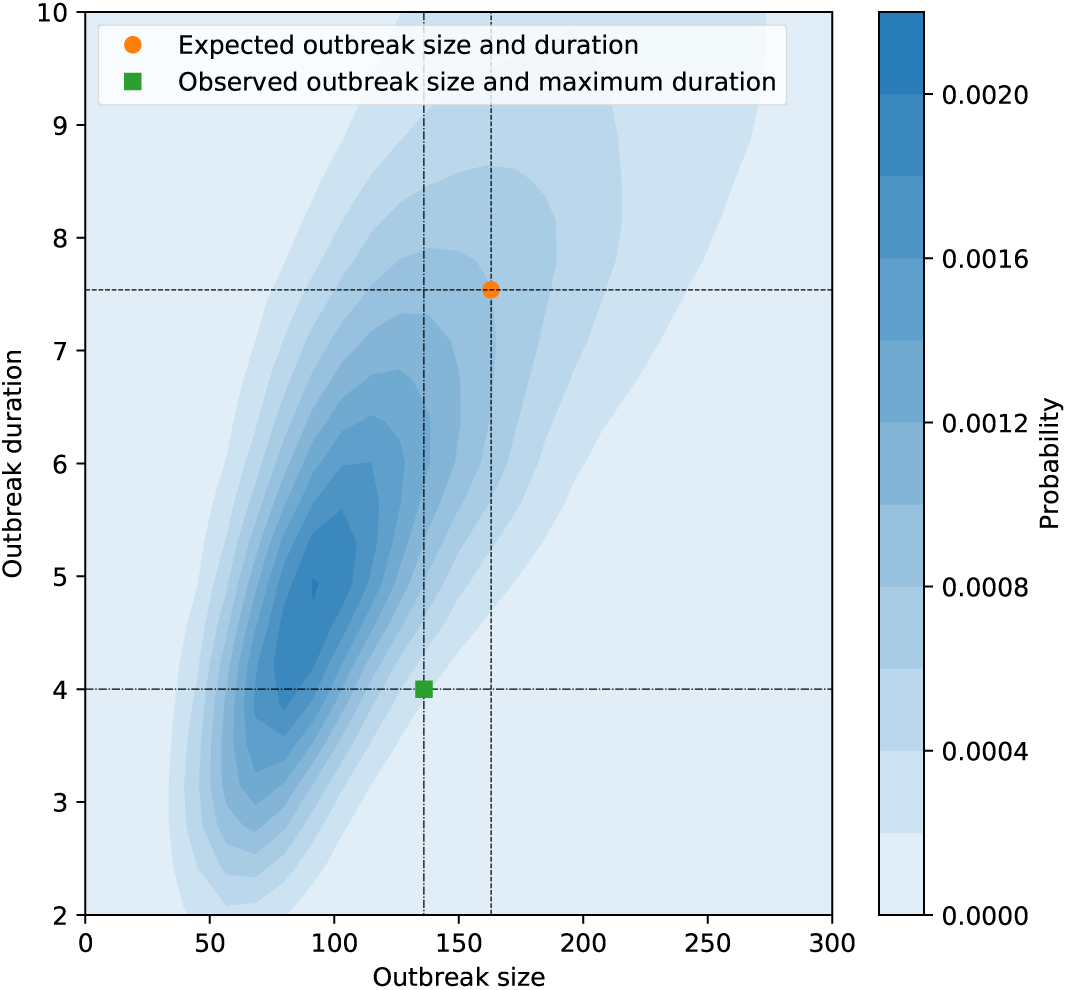
Simulated outbreak size and duration by assuming no control measures. Each simulation was started with 43 infections, and based on reproductive number *R* = 0:74 and dispersion parameter *k* = 0:14 which were estimated from the data collected by February 1st, 2020. The density and mean of duration and outbreak size were estimated based on 5000 Monte-Carlo simulations.

## Discussion

Based on individual-level information of COVID-19 infection cases in the city of Tianjin, we discovered significant transmission heterogeneity (*k* = 0.25, 95%CI: 0.13~0.88) and subcritical transmission potential (*R* = 0.67, 95%CI: 0.54~0.84) and identified one super-spreader who infected 6 individuals, suggesting that the local outbreak in Tianjin was considerably heterogeneous even though it would finally die out. In addition, our numerical results successfully verify the effectiveness of government control deployed on January 28th in Tianjin.

Our finding of significant transmission heterogeneity of COVID-19 outbreak in Tianjin was inconsistent with the previous studies conducted in ShenZhen, China (*k* = 0.58, 95%CI: 0.35~1.18) [13] and in Singapore (*k* = 0.4, 95%CI: 0.1~Inf)[12]. The absence of heterogeneity in these two studies could have been due to the failure of tracking some epidemiological links among cases. In addition, our result was consistent with another study conducted among 46 countries (*k* = 0.1, 95%CI: 0.05~0.2) [14]. However, it should be noted that the large scale spreading of COVID-19 is likely to be heterogeneous because of many extrinsic factors, such as weather [22] and different control measures, may affect the transmission of pathogen, while our confirmation of transmission heterogeneity of COVID-19 in a local outbreak justified that some intrinsic properties of the pathogen might also lead to heterogeneous transmission. Our finding is more meaningful for the development of targeted control measures.

Besides the super spreader in Tianjin as discussed in the results, some other SSEs were also identified based on our criterion in other cities of China, such as in Ningbo, Zhejiang [23], in Harbin, Heilongjiang [24], and in Hongkong [25]. With the tendency of pandemic of COVID-19, all the governments should strengthen controls to prevent SSEs.

Previous studies identified multiple underlying reasons for the emergence of SSEs and high transmission heterogeneity [5]. For example, SSEs of MERS-CoV and SARS-CoV were largely driven by the environmental and clinical factors, such as hospital transfer, substantial delay of diagnosis and fundamentally crowding of population [4, 5]. The transmission heterogeneity and the identified SSE of COVID-19 in Tianjin, however, exhibited different features. The SSE happened in a working place, and there was no obvious delay of diagnosing the super-spreader (3 days from onset of symptom to diagnosis). In addition, the longest transmission chain (as shown in Fig.2) occured after the initiation of Level one response of public health emergency by the Tianjin government. The public were then advised to avoid mass gathering and to stay at home. Obviously, the aforementioned reasons for the transmission heterogeneity in MERS-CoV and SARS-CoV did not hold in the COVID-19 outbreak. Therefore, we suspected the transmission heterogeneity of COVID-19 were driven by some other epidemiological characteristics, such as asymptomatic transmission and contagiousness during incubation period, i.e., transmission can occur before symptom onset. The onset of viral shedding prior to the onset of symptoms, or in cases that remain asymptomatic, is a classic factor that makes infectious disease outbreaks difficult to control [26]. A recent reported transmission chain of large size in Harbin, China were triggered by asymptomatic infections [27], which justified our speculation and highlighted the need for efficient measures such as rapid testing suspected cases to reduce the transmission from subclinical cases.

Additionally, our estimate of transmission potential (*R* < 1) suggested that the spreading of COVID-19 would not cause large infection in Tianjin. This estimate was the average trend over the period of data collection (from January 21st to February 26th), and was considerable lower than that of other studies, especially at the early stage of COVID-19 outbreak [11]. The differences in the reproductive number reported from different studies are largely due to differences methods, differences in data sources and time periods used to estimate the reproductive number. In addition, as indicated with our evaluation based on simulation, the value of *R* < 1 indicated that the control measures imposed in succession by the local government had been considerable reduce the transmissibility of virus.

Regarding the effect of control measure undertaking by the local government, there were changes in point estimates of *R* and *k* of before and after government control, but the difference was not statistically significant with overlapping confidence intervals. Even though, by considering the resulted decrease of 19% infection (from 165 cases to 135 cases) within 25 days, these control measures still had some practical implication. And the non-significant changes in parameter estimates might be due to the short period of observation.

Although the spread of COVID-19 is currently under control in China, it is still facing the risk and challenge brought by the resumption of work and imported cases from other countries. The verification of transmission heterogeneity and SSE in Tianjin outbreak of COVID-19 and the analysis of the underlying reasons remind the government to pay much attention to asymptomatic transmission and other factors that may lead to the transmission heterogeneity and SSEs. Furthermore, more efforts are also needed to explore the emergence of SSE and its contributing factors.

It is also worth mentioning the limitation of our study. Totally 34 imported cases were reported in Tianjin during the period of our study, but we identified 43 transmission chains, which meant some epidemiology links among cases were missing. The estimate of heterogeneity might be driven up if the confirmed cases were condensed into fewer transmission chains. Meanwhile, we only analyzed COVID-19 data from Tianjin (135 confirmed cases), a relatively small sample size compared with the total number in China (exceeds 80,000 confirmed cases), and our case data might be subject to bias or under-reporting, which could not destroy our conclusion of significant heterogeneity [28].

In conclusion, we proved that the transmission of COVID-19 is heterogeneous and identified the existence of one SSE in Tianjin. We also showed that the control measures undertaken by the local government in effect alleviated the outbreak in terms of infection size and duration. As a pandemic which is still spreading worldwide at a startling speed, the transmission characteristics of COVID-19 needs more exploration and investigation in a large scale.

## Data Availability

We retrieved data of the 135 confirmed COVID-19 cases in Tianjin from Tianjin Municipal People’s Government (http://www.tj.gov.cn/xw/ztzl/tjsyqfk/yqtb/). The data that support the findings of this study are available in Github at https://github.com/githublyy0325/COVID-Tianjin-Dataset

## Competing interests

The authors declare that they have no competing interests.

## Author’s contributions

Y.J.Z and X.H.Z designed and directed the project; Y.J.Z developed the theoretical framework; Y.Y.L, L.W, and M.Y.L collected and analyzed the data; Y.J.Z Y.Y.L, L.W, and M.Y.L wrote the article.

## Acknowledgements

The authors acknowledge support from the National Natural Science Foundation of China (grant number 82041023) and Zhejiang University special scientific research fund for COVID-19 prevention and control. The funders had no role in study design, data collection and analysis, decision to publish, or preparation of the manuscript.

## Additional Files

Additional file 1

The additional file 1 is organized as follows. Section A describes detail information of three type of transmission chain. Section B presents the EM algorithm for the estimation of parameters R and k. Section C gives technical details for the construction of bootstrap confidence interval (CI) of the estimates from EM algorithm

Additional file 2 — Data

The data information includes chain size, chain type, the Number of primary case, ID, onset of symptoms of primary case, confirmed date of primary case.

## Notes

### Competing Interest Statement

The authors have declared no competing interest.

## References

1. (WHO), W.H.O., et al.: Statement on the second meeting of the international health regulations (2005) emergency committee regarding the outbreak of novel coronavirus (2019-ncov). geneva: Who; 30 jan 2020.[accessed 01 feb 2020]. Geneva, Switzerland (2005)

2. Lloyd-Smith, J.O., Schreiber, S.J., Kopp, P.E., Getz, W.M.: Superspreading and the effect of individual variation on disease emergence. Nature 438(7066), 355–359 (2005)

3. Kucharski, A., Althaus, C.: The role of superspreading in middle east respiratory syndrome coronavirus (mers-cov) transmission. Euro surveillance 20(25), 21167 (2015)

4. Wong, G., Liu, W., Liu, Y., Zhou, B., Bi, Y., Gao, G.F.: Mers, sars, and ebola: The role of super-spreaders in infectious disease. Cell Host & Microbe 18(4), 398–401 (2015)

5. Stein, R.A.: Super-spreaders in infectious diseases. International Journal of Infectious Diseases 15(8), 510–513 (2011)

6. Kucharski, A.J., Althaus, C.L.: The role of superspreading in middle east respiratory syndrome coronavirus (mers-cov) transmission. Eurosurveillance 20(25), 21167 (2015)

7. Lau, M.S.Y., Dalziel, B.D., Funk, S., McClelland, A., Tiffany, A., Riley, S., Metcalf, C.J.E., Grenfell, B.T.: Spatial and temporal dynamics of superspreading events in the 2014–2015 west africa ebola epidemic. Proceedings of the National Academy of Sciences of the United States of America 114(9), 2337–2342 (2017)

8. Faye, O., Boëlle, P.-Y., Heleze, E., Faye, O., Loucoubar, C., Magassouba, N., Soropogui, B., Keita, S., Gakou, T., Bah, E.H.I., Koivogui, L., Sall, A.A., Cauchemez, S.: Chains of transmission and control of ebola virus disease in conakry, guinea, in 2014: an observational study. Lancet Infectious Diseases 15(3), 320–326 (2015)

9. Li, Q., Guan, X., Wu, P., Wang, X., Zhou, L., Tong, Y., Ren, R., Leung, K.S., Lau, E.H., Wong, J.Y., et al.: Early transmission dynamics in wuhan, china, of novel coronavirus–infected pneumonia. New England Journal of Medicine (2020)

10. Riou, J., Althaus, C.L.: Pattern of early human-to-human transmission of wuhan 2019 novel coronavirus (2019-ncov), december 2019 to january 2020. Eurosurveillance 25(4) (2020)

11. Liu, Y., Gayle, A.A., Wilder-Smith, A., Rocklöv, J.: The reproductive number of covid-19 is higher compared to sars coronavirus. Journal of Travel Medicine (2020)

12. Tariq, A., Lee, Y., Roosa, K., Blumberg, S., Yan, P., Ma, S., Chowell, G.: Real-time monitoring the transmission potential of covid-19 in singapore, february 2020. medRxiv (2020). doi:10.1101/2020.02.21.20026435. https://www.medrxiv.org/content/early/2020/03/12/2020.02.21.20026435.full.pdf

13. Bi, Q., Wu, Y., Mei, S., Ye, C., Zou, X., Zhang, Z., Liu, X., Wei, L., Truelove, S.A., Zhang, T., et al.: Epidemiology and transmission of covid-19 in shenzhen china: Analysis of 391 cases and 1,286 of their close contacts. MedRxiv (2020)

14. Endo, A., Abbott, S., Kucharski, A.J., Funk, S., et al.: Estimating the overdispersion in covid-19 transmission using outbreak sizes outside china. Wellcome Open Research 5(67), 67 (2020)

15. Liu, Y., Eggo, R.M., Kucharski, A.J.: Secondary attack rate and superspreading events for sars-cov-2. The Lancet 395(10227), 47 (2020)

16. Wu, W., Li, Y., Wei, Z., Zhou, P., Lyu, L., Zhang, G., Zhao, Y., He, H., Li, X., Gao, L., et al.: Investigation and analysis on characteristics of a cluster of covid-19 associated with exposure in a department store in tianjin. Zhonghua liu Xing Bing xue za zhi= Zhonghua Liuxingbingxue Zazhi 41(4), 489–493 (2020)

17. Blumberg, S., Lloyd-Smith, J.O.: Inference of r 0 and transmission heterogeneity from the size distribution of stuttering chains. PLoS Computational Biology 9(5) (2013)

18. The Proposal of New Coronavirus Pneumonia Prevention(Sixth Edition). http://www.chinacdc.cn/jkzt/crb/zl/szkb_11803/jszl_11815/202003/W020200309376009304000.pdf

19. Tianjin Municipal People’s Government. http://www.tj.gov.cn/xw/ztzl/tjsyqfk/yqtb/

20. Tianjin: Come up with 18 Policies for Epidemic Prevention and Control. http://www.tj.gov.cn/xw/spxw/202001/t20200129_3668285.html

21. Lauer, S.A., Grantz, K.H., Bi, Q., Jones, F.K., Zheng, Q., Meredith, H.R., Azman, A.S., Reich, N.G., Lessler, J.: The incubation period of coronavirus disease 2019 (covid-19) from publicly reported confirmed cases: Estimation and application. Annals of Internal Medicine

22. Neher, R.A., Dyrdak, R., Druelle, V., Hodcroft, E.B., Albert, J.: Potential impact of seasonal forcing on a sars-cov-2 pandemic. Swiss Medical Weekly 150(1112) (2020)

23. Health Commission of Ningbo. http://wjw.ningbo.gov.cn/col/col142/index.html

24. All 9 Family Members in New Year’s Eve Dinners in Heilongjiang Were Confirmed. http://hlj.people.com.cn/GB/n2/2020/0205/c220024-33767665.html

25. Nine Members of Hongkong Family Feared Infected After Sharing Hotpot. https://www.straitstimes.com/asia/east-asia/coronavirus-nine-members-of-hong-kong-family-feared-infected-after-sharing-hotpot

26. Fraser, C., Riley, S., Anderson, R.M., Ferguson, N.M.: Factors that make an infectious disease outbreak controllable. Proceedings of the National Academy of Sciences of the United States of America 101(16), 6146–6151 (2004)

27. Health Commission of Heilongjiang Province. http://yiqing.ljjk.org.cn/index/patients/newsinfo/id/1887.html

28. Lloyd-Smith, J.O.: Maximum likelihood estimation of the negative binomial dispersion parameter for highly overdispersed data, with applications to infectious diseases. PloS one 2(2) (2007)

